# An artificial intelligence system for predicting mortality in COVID-19 patients using chest X-rays: a retrospective study

**DOI:** 10.1101/2021.09.22.21263956

**Authors:** Avinash Nanivadekar, Kapil Zirpe, Ashutosh Dwivedi, Rajan Patel, Richa Pant, Tanveer Gupte, Rohit Lokwani, Dinesh Shende, Viraj Kulkarni, Amit Kharat

## Abstract

**Background:** Early prediction of disease severity in COVID-19 patients is essential. Chest X-ray (CXR) is a faster, widely available, and less expensive imaging modality that may be useful in predicting mortality in COVID-19 patients. Artificial Intelligence (AI) may help expedite CXR reading times, and improve mortality prediction. We sought to develop and assess an artificial intelligence system that used chest X-rays and clinical parameters to predict mortality in COVID-19 patients.

**Methods:** A retrospective study was conducted in Ruby Hall Clinic, Pune, India. The study included patients who had a positive real-time reverse transcriptase-polymerase chain reaction (RT-PCR) test for COVID-19 and at least one available chest X-ray at the time of their initial presentation or admission. Features from CXR images and clinical parameters were used to train the Random Forest model.

**Results:** Clinical data from a total of 201 patients was assessed retrospectively. The average age of the cohort was 51.4±14.8 years, with 29.4% of the patients being over the age of 60. The model, which used CXRs and clinical parameters as inputs, had a sensitivity of 0.83 [95% CI: 0.7, 0.95] and a specificity of 0.7 [95% CI: 0.64, 0.77]. The area under the curve (AUC) on receiver operating characteristics (ROC) was increased from 0.74 [95% CI: 0.67, 0.8] to 0.79 [95% CI: 0.72, 0.85] when the model included features of CXRs in addition to clinical parameters.

**Conclusion:** An Artificial Intelligence (AI) model based on CXRs and clinical parameters demonstrated high sensitivity and can be used as a rapid and reliable tool for COVID-19 mortality prediction.

## INTRODUCTION

The novel human Coronavirus disease 2019 (COVID-19) was first reported in Wuhan, Hubei province of China on December 31, 2019, and has eventually spread worldwide (1). As of 14 September 2021, more than 224 million confirmed cases of COVID-19 had been reported worldwide (2). India is one of the worst affected countries, with over 33 million positive cases and more than 4.41 lakh deaths reported till the first week of September 2021 (3). There has been an extraordinary global health response in an effort to reduce transmission and mortality due to COVID-19 (4). In order to optimize hospital resources, health-care providers must efficiently triage patients based on the severity of COVID-19 infection. An early prognostication of disease severity can facilitate timely allocation of resources to patients who are critically ill or are likely to advance to a critical stage and require intensive care. Real-time reverse transcriptase-polymerase chain reaction (RT-PCR) is considered the gold standard for COVID-19 diagnosis. However, the high false negative rate and long turnaround time limit its effectiveness, allowing the infection to spread within communities (5). Additionally, the limited testing infrastructure for RT-PCR and long turnaround time are major concerns in confirming the COVID-19 diagnosis in early stages in India.

Medical imaging is one of the most feasible methods for diagnosing and predicting the severity of COVID-19. Chest Computed Tomography (CT) is routinely used for early diagnosis and prognosis of COVID-19 in patients (6-7). Although CT has high sensitivity, its utility is constrained due to limited availability at primary care settings, high patient volumes, portability and the difficulty of transporting oxygen-dependent critically ill COVID-19 patients. On the contrary, chest X-ray has relatively better availability, low cost, and portability, making it a potential first-line triage tool in resource poor settings (8). Chest X-rays are being utilized in the context of COVID-19 to determine the likelihood of hospital admission, duration of hospital treatment, and risk of critical outcomes (9-11).

During the current COVID-19 pandemic, there was a sudden surge in demand for diagnostic tools, exposing the lacunae in diagnostic infrastructure and reporting methods. There have also been instances of discrepancies in radiologists’ interpretations and diagnostic errors (12). Deep learning, a popular research area of Artificial Intelligence (AI), may help accelerate CXR reading times, making it an excellent candidate in the clinical delivery of patient care. Utilizing artificial intelligence, Chest X-rays can strengthen clinical data in predicting the severity of COVID-19 in patients (13-14).

A recent study by Murphy *et al*. reported that an AI system could detect COVID-19 pneumonia on CXRs with the same accuracy as six independent radiologists (15). However, the role of AI as a prognostic tool using CXRs in COVID-19 patients has yet to be explored in detail. Only a few studies have integrated artificial intelligence into chest X-ray analysis to predict progression risk, demonstrating the advantage of chest X-rays over the clinical data-only prediction models (16-17).

Herein, we aimed to evaluate the association between CXR findings and clinical outcomes, as well as the use of an AI system as an early prognostic tool in detecting COVID-19. The study intends to introduce an AI based prognosis model that takes an X-Ray image and other clinical data as input and predicts the outcome to categorize the patient as high risk (death) or low-risk (discharge). This is significant because the outcome of COVID-19 is heavily influenced by patient demographics, as well as some key clinical factors.

## MATERIALS AND METHODS

### Clinical data

The study was conducted at Ruby Hall clinic, a tertiary care centre in Pune, Maharashtra, India. The data used for AI modeling included radiological findings on Chest X-rays (CXRs), and clinical findings and observations such as symptoms and comorbidities. The details of all the clinical and radiological parameters of the patients diagnosed with COVID-19 along with the patients’ information have been provided in Supplementary Table 1. Out of all the different parameters shown in Supplementary table 1, only 10 significant parameters were used to build an AI model. The study was approved by the institutional ethics committee of the hospital and the patients’ identities were concealed throughout all the steps of data handling.

### Participants

The study was conducted at Ruby Hall Clinic, Pune, and no filters were used based on the demographic/geospatial or contact tracing information of the patient. In this retrospective investigation, only patients with a positive RT-PCR report and at least one X-ray imaging post-admission were included. The study did not include the patients’ past and current hospital treatment procedures.

### AI Model training

Random Forest model was trained on the selected clinical and demographic parameters. Random forests are learning methods for classification tasks that operate by constructing a group of decision trees at training time, which helps in decision making. The end-to-end process involves pre-processing the data that involves converting raw data to data suitable for the machine learning model. For this study, the data was cleaned (details in the data pre-processing section) and the clinical parameters were reduced to the significant ones. Hyper-parameter grid search was used for optimization to find the right parameters to get the best model performance. Hyper-parameters are the parameters that are not directly learnt by the algorithm and are tuned manually. If an optimal combination of hyper-parameters is set for the model, it optimizes better for specific metrics. Grid search optimization essentially provides a range of hyper-parameters to the model, which then attempts to find the optimal set by cross-validating on the entire dataset by attempting all possible combinations in different iterations of the algorithm. The threshold of 0.4 was used to classify the patient as being at high risk of mortality using the optimization algorithm and cross-validation on the entire dataset.

### Data pre-processing

This involves two steps: data cleaning and dimensionality reduction. Data was anonymized for any of the patient’s personal or demographic details, apart from the patient’s age and gender, as well as the date of the scan that were used for modeling. Entries for patients with missing or null values were removed. Due to skewness and the sensitivity of medical data, statistical techniques were not applied to fill up the missing entries. After removing the entries, separate features for symptoms were created, and manually pre-processed for human errors. Every symptom was treated as a separate parameter. All the binary entries for presence/absence of symptoms were replaced with boolean values (0/1). The co-morbidity column was also converted to boolean values, which represent their presence or absence. The absolute difference between the date of onset of symptoms and the scan date was calculated and added as a feature. Clinically, if the patient has COVID-19 pneumonia in the lungs, it is likely to worsen if not treated promptly. Hence, the number of days between the X-ray and the onset of symptoms would presumably be useful in determining the outcome.

We further added two features from the X-ray COVID-19 detection model: probability of prediction and area of the predicted mask with respect to the image size. The probability of prediction indicates the confidence with which the model has predicted COVID-19 in the X-rays. The area of the mask showed the severity of the COVID-19 pneumonia in the lungs. Both of these features were normalized to values in the range [0,100], where 0 = no lung involvement, and 100 = severe lung involvement. To avoid any discrepancies, we removed the patients with “admitted” as their outcome and kept a boolean output, where discharge was assigned 0 and death was assigned 1. The final data used for model building was that of 201 patients, with the prevalence of death being 8.9%. The number of parameters was reduced from 66 to 32 features using the above filtering and pre-processing criteria. The parameters were then verified for accuracy before training the model.

### Random Forest Model

We trained the Random Forest model on the dataset. Hyper-parameter grid search optimization was used on the whole set to finalize the algorithm’s hyper-parameters. The algorithm was trained with Gini as the entropy, with a total of 100 estimators and a maximum depth of 2. Class weights were used to balance and penalize the classes based on the data skewness. We performed four-fold cross-validation on the entire set before calculating the aggregated results.

The flowchart depicting the prediction of COVID-19 associated mortality by the model is represented in Figure 1.

**Figure 1:**
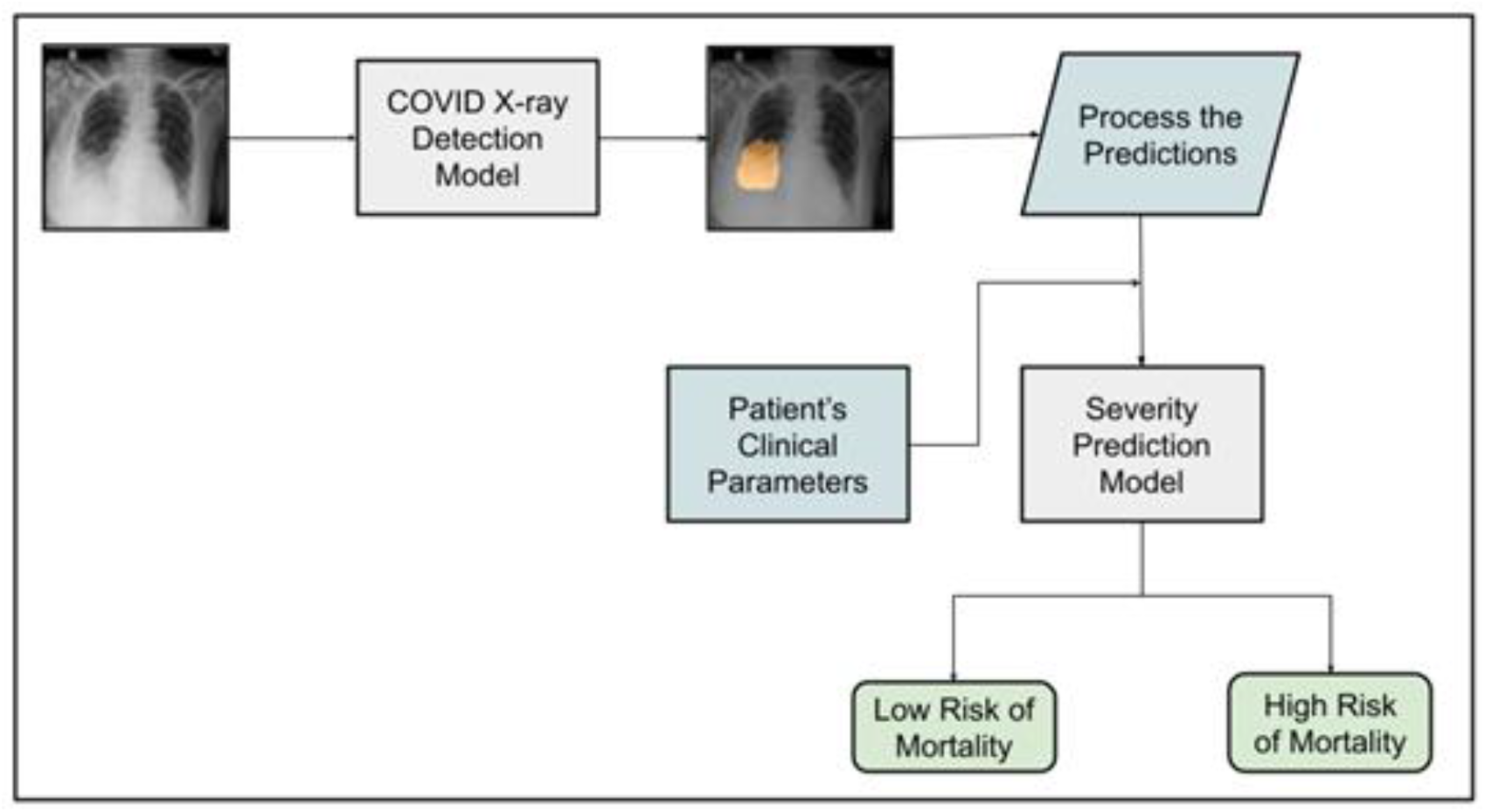
Flowchart depicting the process of severity prediction by the AI model.

### Statistical analysis

McNemar’s test was used to compute the p-value for each demographic feature between patient subgroups. McNemar’s test was also used to calculate the p value between survivors and non-survivors for each radiographic finding on the CXRs. p value < 0.05 was considered statistically significant. The 95% Confidence Interval (CI) was calculated using the Empirical Bootstrapping method. To assess the performance of the AI model, a receiver operating characteristic (ROC) curve was generated. The area under the ROC curve with 95% CI was calculated.

### RESULTS AND OBSERVATIONS

Clinical data from 201 patients was analyzed retrospectively. There was a male preponderance (74.1% males vs. 25.9% females) among the patients, but there was no significant difference in the mortality rate between the two genders. The average age of the cohort was observed to be 51.4 ± 14.8 years, with 29.4 % of the patients being over the age of 60. There was a highly significant difference in mortality proportion (78.8% vs. 22.2%) between age groups > 60 and ≤60 years, respectively (p<0.0001). The vast majority of patients (60.2%) had associated comorbidities, and the proportion of cases with comorbidities in the mortality group was significantly higher (88.9% vs. 11.1%) than their non-comorbid counterparts (p=0.01). The demographic characteristics of the participants along with the clinical outcome are provided in Table 1.

**Table 1:** Demographic characteristics of the study participants.

| Demographic Characteristics | Subgroups | Parameters | Outcome |  | Total | p value |
| --- | --- | --- | --- | --- | --- | --- |
|  |  |  | Survivors<br>(N=183) | Deceased<br>(N=18) |  |  |
| Gender | Females | Number | 46 | 6 | 52 | 0.414 |
|  |  | % | 25.1% | 33.3% | 25.9% |  |
|  | Males | Number | 137 | 12 | 149 |  |
|  |  | % | 74.9% | 66.7% | 74.1% |  |
| Age groups | $\leq 60$ years | Number | 138 | 4 | 142 | <0.0001 |
|  |  | % | 75.4% | 22.2% | 70.6% |  |
|  | > 60 years | Number | 45 | 14 | 59 |  |
|  |  | % | 24.6% | 77.8% | 29.4% |  |
| Comorbidity | Absent | Number | 78 | 2 | 80 | 0.01 |
|  |  | % | 42.6% | 11.1% | 39.8% |  |
|  | present | Number | 105 | 16 | 121 |  |
|  |  | % | 57.4% | 88.9% | 60.2% |  |

201 CXRs were acquired in the frontal position. According to the radiographic findings, septal thickening was observed in 81.5% of the CXRs, and consolidation was found in 61.7% of CXRs. Vascular thickening was present in 38% CXRs, whereas pleural effusion was found in only 5.5% CXRs. While the presence of these radiographic features was comparable between survivors and non-survivors, the proportion of pleural effusion in non-survivors was significantly higher than in survivors (27.8% versus 3.3%). Different radiographic findings among survivors and non-survivors are listed in Table 2.

**Table 2:** Comparison of radiographic findings among survivors and non-survivors.

| Radiographic Finding |  | Parameter | Outcome |  | Total | p value |
| --- | --- | --- | --- | --- | --- | --- |
|  |  |  | Survivors<br>(N=183) | Deceased<br>(N=18) |  |  |
| Pleural Effusion | Absent | Number | 177 | 13 | 190 | 0.001 |
|  |  | % | 96.7% | 72.2% | 94.5% |  |
|  | Present | Number | 6 | 5 | 11 |  |
|  |  | % | 3.3% | 27.8% | 5.5% |  |
| Vascular thickening | Absent | Number | 114 | 10 | 124 | 0.614 |
|  |  | % | 62.6% | 55.6% | 62.0% |  |
|  | Present | Number | 68 | 8 | 76 |  |
|  |  | % | 37.4% | 44.4% | 38.0% |  |
| Septal thickening | Absent | Number | 33 | 4 | 37 | 0.751 |
|  |  | % | 18.1% | 22.2% | 18.5% |  |
|  | Present | Number | 149 | 14 | 163 |  |
|  |  | % | 81.9% | 77.8% | 81.5% |  |
| Consolidation | Absent | Number | 72 | 5 | 77 | 0.242 |
|  |  | % | 39.3% | 27.8% | 38.3% |  |
|  | Present | Number | 111 | 13 | 124 |  |
|  |  | % | 60.7% | 72.2% | 61.7% |  |

The clinical data-based severity prediction of the model had an F1 score of 0.26 [95% CI: 0.18, 0.35] and a kappa score of 0.17 [95% CI: 0.1, 0.23]. The algorithm produced a sensitivity of 0.83 [95% CI: 0.7, 0.95] and a specificity of 0.61 [95% CI: 0.56, 0.66]. When CXRs were also included along with the clinical parameters, the specificity was increased to 0.7 [95% CI: 0.64, 0.77] from 0.61 [95% CI: 0.56, 0.66]. The F1 score and the kappa score were increased from 0.26 [95% CI: 0.18, 0.35] and 0.17 [95% CI: 0.1, 0.23] to 0.34 [0.21,0.47] and 0.24 [0.12, 0.35], respectively. The specificity (discharge-recall) was also increased from 0.61 [0.56, 0.66] to 0.7 [0.64, 0.77]. The crucial features included the age, probability, and area of infection predicted by the X-ray AI model. The performance of the AI model is depicted in Table 3.

**Table 3:** Performance of the AI algorithm with and without X-rays.

| **Metric** | **Values for AI with X-rays**<br>**[95% CI]** | **Values for AI without X-rays**<br>**[95% CI]** |
| --- | --- | --- |
| Sensitivity (Death) | 0.83 [0.7, 0.95] | 0.83 [0.7, 0.95] |
| Precision (Death) | 0.22 [0.12, 0.32] | 0.15 [0.1, 0.22] |
| Specificity (Discharge-Recall) | 0.7 [0.64, 0.77] | 0.61 [0.56, 0.66] |
| F1-score(Death) | 0.34 [0.21, 0.47] | 0.26 [0.18,0.35] |
| Kappa Score | 0.24 [0.12, 0.35] | 0.17 [0.1,0.23] |

Incorporation of CXRs in addition to the clinical parameters augmented the data in predicting mortality in COVID-19 patients. The receiver operating characteristic (ROC) curve that illustrates mortality prediction had an area under the curve (AUC) of 0.74 [0.67, 0.80], which was increased to 0.80 [0.72, 0.85] when the model also included features of CXRs (Figure 2).

**Figure 2:**
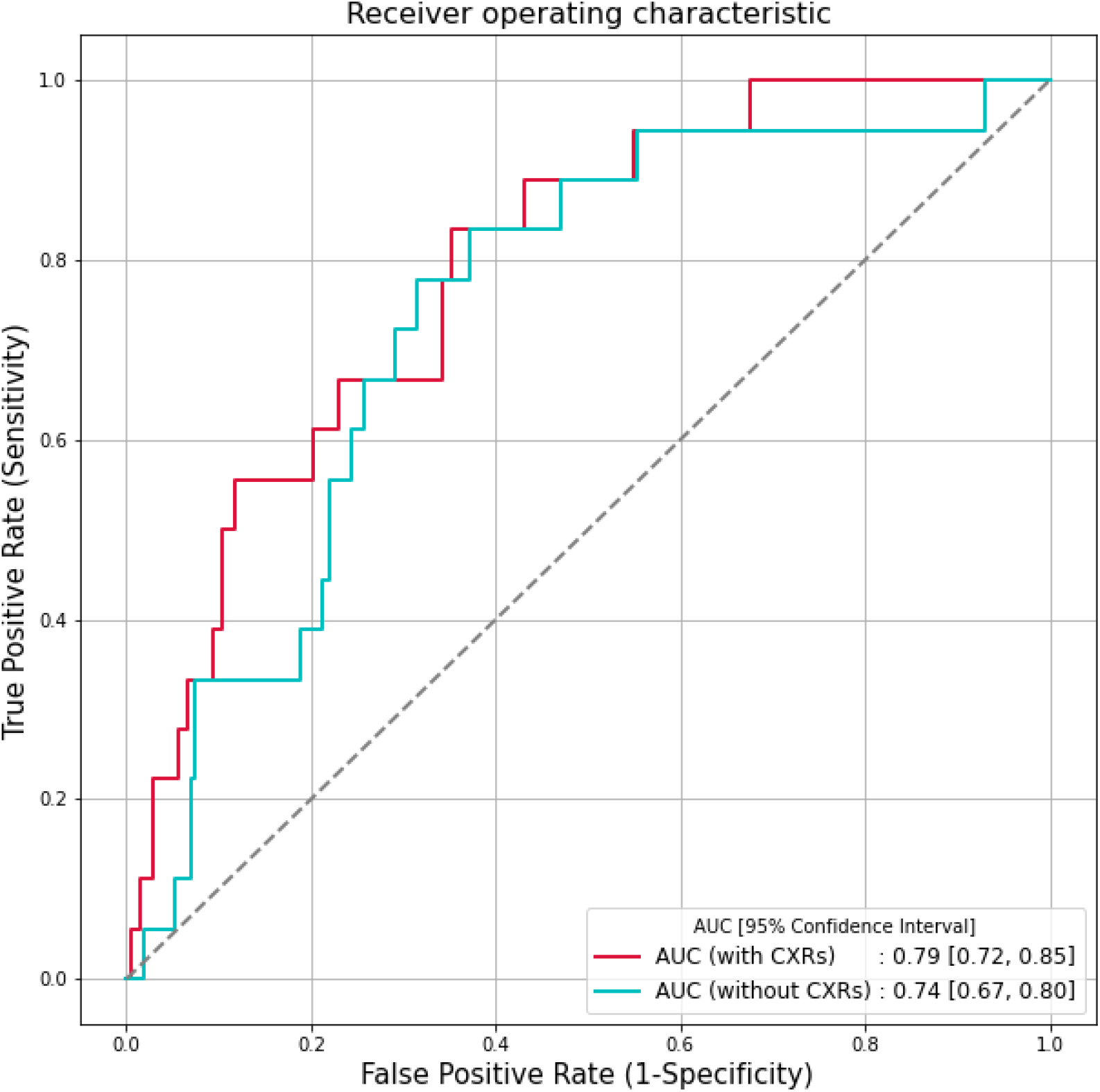
Receiver operating characteristic (ROC) plots for COVID-19 mortality prediction with and without CXRs as a parameter.

The Random forest model was trained as described in materials and methods. Each variable was given an importance rank according to the tree statistics of the training set. The age of the patient was ranked among the top predictors of mortality in the cohort. This was followed by the AI prediction of the CXRs and co-morbidities amongst the patients. Different presenting symptoms within the cohort, such as dyspnea, vomiting, cough, and diarrhoea, were given boolean values, i.e., 0 for the absence and 1 for the presence of a particular symptom. The top 10 independent clinical variables associated with mortality are listed in Table 4. These clinical parameters describe the feature importance or dominance necessary to understand the decision-making process. A graphical representation of these features along with their percentage of importance is shown in Supplementary Figure 1.

**Table 4:** Features used in AI decision-making and their description

| Importance Rank | Feature Name | Feature Description |
| --- | --- | --- |
| 1 | Age | Age of the Patient |
| 2 | X-ray AI Prediction-Confidence | Probability of prediction [0, 100] |
| 3 | X-ray AI Prediction-Area of the mask | Percentage area of the image covered by the predicted mask [0, 100] |
| 4 | Comorbidities | Includes Hypertension, Diabetes, Hypothyroidism, Vertigo, and Asthma amongst others. The column is Boolean: if any of these comorbidities exist, it's marked as 1 otherwise 0 |
| 5 | Days (Date of Scan- Date of Symptoms) | Absolute Difference of the date of CXR and the date of onset of symptoms |
| 6 | Clinical condition of patient at the time of admission (stable/serious) | Condition of the patient at time of admission. The column is Boolean: if patient was stable, it's marked as 0 otherwise 1 |
| 7 | Dyspnea | Presenting Symptom - Marked 0 for absence and 1 for presence |
| 8 | Vomiting | Presenting Symptom - Marked 0 for absence and 1 for presence |
| 9 | Cough | Presenting Symptom - Marked 0 for absence and 1 for presence |
| 10 | Diarrhoea | Presenting Symptom - Marked 0 for absence and 1 for presence |

## DISCUSSION

The ongoing COVID-19 catastrophe has created an unprecedented burden on the Indian healthcare infrastructure (18). It is one of a kind situation that necessitates rapid detection of disease severity in order to reduce mortality rates through early interventions (19-20). Chest X-rays are a useful imaging modality that has helped with diagnosis and prognosis of COVID-19. The value of including chest X-rays to clinical data for disease prognosis, on the other hand, needs to be thoroughly investigated. An artificial intelligence model based on the initial chest X-rays and clinical data of COVID-19 patients presented to a tertiary care hospital was used in this study and was shown to significantly improve prognostic ability. Additionally, the model based on deep-learning features extracted from chest X-rays, was found to be an effective tool for improving the utility of radiologist-interpreted X-rays in COVID-19 diagnosis and severity prediction.

We observed that the radiographic features in our cohort were consistent with previous reports (9, 21-22). The distribution of lung opacities, consolidation (61.7%) and septal thickening were typically bilateral and very few cases of pleural effusion (5.5%) were observed. The findings are consistent with previous studies that have validated the use of initial CXR severity scores as independent outcome predictors (7–9). The study has reiterated the finding that COVID-19 severity, assessed on the CXRs at primary presentation, is a valuable prognostic factor that should be considered by medical practitioners when making triage decisions. Our AI model had a sensitivity of 83% [95% CI: 0.7, 0.95] and specificity of 70% [95% CI: 0.64, 0.77] that was comparable to the AI model described by Mushtaque *et al*. (23), which had sensitivity of 79% and specificity of 58.8%. A recent study by Jiao *et al*. reported that when chest X-rays were added to clinical data for severity prediction, the area under the receiver operating characteristic (AUC-ROC) curve increased from 0.821 [95% CI: 0.796, 0.828] to 0·846 [95% CI: 0.815, 0.852] on internal testing and 0·731 [95% CI: 0.712, 0.738] to 0·792 [95% CI: 0.780, 0.803] on external testing (13). Similarly, in our study, it was observed that the area under the receiver operating characteristic (AUC-ROC) curve for mortality prediction increased from 0.74 [95% CI: 0.67, 0.8] to 0.79 [95% CI: 0.72, 0.85] when the model included features of X-rays in addition to clinical features. Additionally, the F1 score was increased from 0.26 [95% CI: 0.18, 0.35] to 0.34 [95% CI: 0.21, 0.47] when AI-calculated X-ray parameters were added.

According to an Ernst & Young Intelligent Privacy Automation (EY-IPA) study, the domestic telemedicine market is expected to reach USD 5.5 billion by 2025, and the Indian healthcare industry will need to transition from traditional in-person doctor-patient interaction to digitized remote consultations (24). AI-enabled telehealth contributes to quality improvement and enhancement of existing processes, as well as the implementation of new models of care (25-26), which are desperately needed in the overburdened Indian healthcare system.

This study established a substantial role of AI in diagnostic radiology, and further implementation of AI in clinical imaging is poised to transform the practice of radiology.

## CONCLUSION

A relatively high sensitivity of our model makes it a rapid and reliable tool for COVID-19 mortality prediction, particularly in resource-constrained environments. Such AI models have the potential to be used as an initial screening tool for triaging COVID-19 patients, as well as to increase vigilance for severe cases. The usefulness of the model was successfully demonstrated in a tertiary care centre in India for prediction of COVID-19 mortality and can be easily adopted by imaging experts and clinicians.

However, since the study is retrospective, it may result in an observer’s bias. Because this was a single-centre study conducted in one of the referral tertiary care centres in India, it’s likely that only moderate to severe cases were included. Further studies with multicentric data may reduce the selection bias and improve the sensitivity of the model with greater precision.

## Data Availability

The data supporting the findings of this study are available within the article and supplementary materials.

