## Supplementary file for "An artificial intelligence system for predicting mortality in COVID-19 patients using chest X-rays: a retrospective study"

### Supplementary data

**Supplementary Table 1:** Clinical and radiological parameters of the patients diagnosed with COVID-19.

| Patient's Personal Information | CT and CXR Findings | Hospital Data | Clinical History | Travel History | Transmission History | Testing History | Current Status |
|---|---|---|---|---|---|---|---|
| 1. PID<br>2. Name (RHC)<br>3. Gender (RHC)<br>4. Age (RHC)<br>5. Name (Registered with PMC)<br>6. Age (Registered with PMC)<br>7. Sex (Registered with PMC)<br>8. Address (Detail address of patient)<br>9. Contact no.<br>10. Occupation<br>11. District/ Corporation (by place of residence)<br>12. State (by patients Residence)<br>13. Nationality | 14. HRCT Score<br>15. Date of Scan<br><br>Patterns<br>16. GGO,<br>17.Consolidation,<br>18. Septal thickening,<br>19. Vascular thickening,<br>20. Pleural Effusion<br><br>Location<br>21. RUL<br>22. RML<br>23. RLL<br>24. LUL<br>25. LLL | 26. Case No.<br>27. District<br>28. GoI COVID portal (SSSNO)<br>29. District / Corporation case number<br>30. Sr. No. | 31. Name of 1st admitted Hospital<br>32.District/ Corporation of Admitted Hospital<br>33. Date of 1st admission<br>34.Symptoms at the time of 1st admission<br>35. Date of onset of symptoms<br>36. Presenting symptoms<br>37. Comorbidity condition<br>38. Clinical condition at time of admission<br>39. Treatment Protocol<br>40. Name of 2nd Hospital<br>41. Date of admission in 2nd Hospital | 42. Travel history outside India<br>43. If yes Name of Visited Country<br>44. Travel within India (Yes/No)<br>45. If yes Name of City<br>46. Attended mass gathering (Yes/No)<br>47. If yes, Place & Address of mass gathering | 48. COVID-19 +ve family member<br>49. H/o contact with COVID-19 +ve person<br>50. Total Number of Contacts<br>51. No. of contacts admitted<br>52. No. of contacts at Home<br>53. No. of samples of contacts sent<br>54. No. of contacts found Positive | 55. Date of 1st Sample Collection<br>56. sample collection centre<br>57. Sample tested at<br>58. Date of result of 1st sample<br>59. Day -14 Test Date<br>60. Day-14 Test Result<br>61. Day-15 Test Date<br>62. Day-15 Test Result | 63. Ward<br>64.Condition today<br>65. Swab Status<br>66. Outcome |

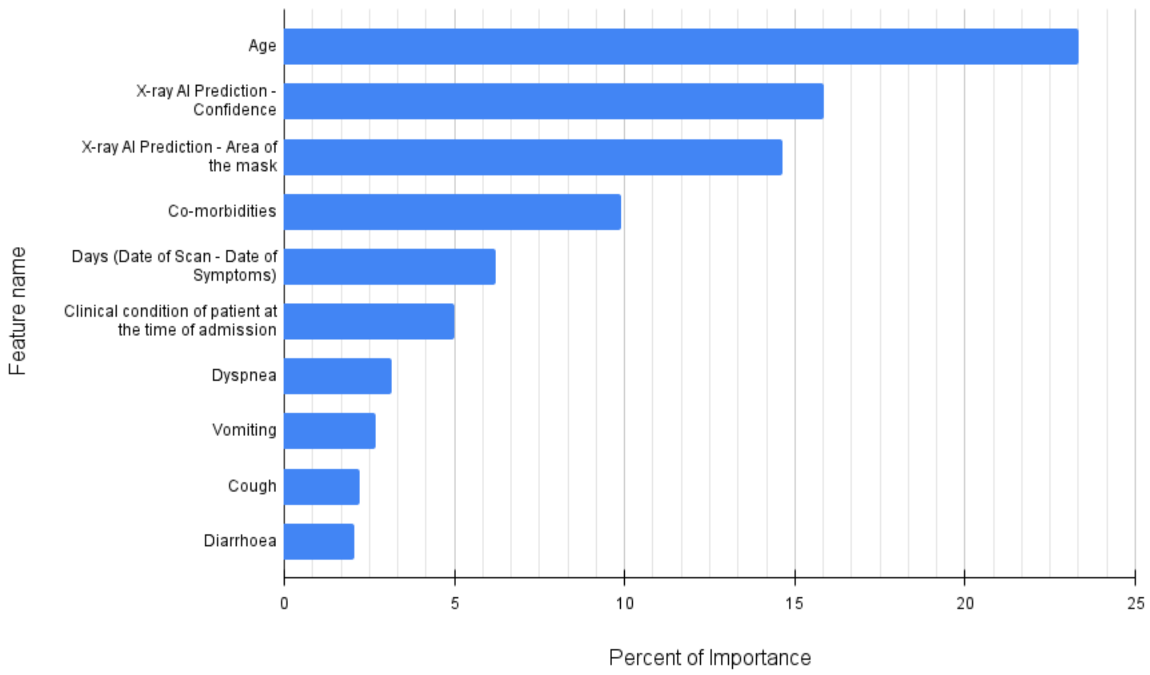


**Supplementary Figure 1: Random Forest model ranking top 10 clinical variables for predicting mortality associated with COVID-19.** The Y-axis indicates the clinical features arranged according to their rank of importance. The X-axis indicates their percent of importance.
